# Management of COVID-19-related Arterial Thrombosis Leading to Acute Limb-threatening Ischemia

**DOI:** 10.1101/2021.03.20.21252888

**Authors:** Omer Ehsan, Mohammad Iqbal Khan, Muhammad Waqas Raza

## Abstract

**Objective:** Examine the occurrence and clinical outcomes of arterial thrombosis leading to limb-threatening ischemia in patients with coronavirus-2019 (COVID-19).

**Study design:** Prospective, descriptive case series.

**Patients and Methods:** Forty-four patients with COVID-19 infection presenting with critical limb ischemia were managed between March 2020 and December 2020. The patients were divided into three groups based on the mode of presentation: 1) those who had been admitted; 2) those presenting in the emergency department; and 3) those presenting to vascular clinics. Clinical evidence suggesting limb ischemia was evaluated with computerized tomographic angiogram. Vascular care was designed according to the need of individual patients, through anticoagulation, revascularization by thrombo-embolectomy, or bypass grafting and amputation.

**Results:** Ten major amputations and 4 deaths (all in patients admitted) occurred among the 44 patients (9.1%). Most patients (32/44) were males, mean age was 55, and limb ischemia occurred among patients as young as 29. The initial period of ischemia was often not appreciated by patients and physicians. Critical limb ischemia was often not correlated with the severity of COVD symptoms: of 17 patients who presented through the emergency room with limb-threatening ischemia, 10 (58.9%) were asymptomatic for respiratory and general symptoms. Comorbidities were common among all 3 patient groups (26/44; 59%). Anticoagulants did not consistently prevent thromboembolic events since all admitted patients were receiving low molecular weight heparin. The rate of revascularization was lower in this population than in the general population with similar limb ischemia.

**Conclusion:** Acute limb ischemia in patients with COVID-19 is a vascular emergency that can result in limb loss and even death. The severity of respiratory infection and other symptoms of COVID often are not consistent with the severity and level of vascular involvement. Timely recognition and tailored intervention is needed to save limbs in this population.

**What this paper adds to existing knowledge:** COVID-19 infection predisposes to arterial and venous thrombosis, but data on arterial thrombosis are sparse. This report describes a series of 44 COVID-19 patients who had arterial thrombosis causing limb-threatening ischemia. Most noteworthy characteristics of the condition are its severity (10 major amputations and 4 deaths); subtle initial expression of the ischemia, which may lead to delayed diagnosis; severity and level of vascular involvement poorly correlated with respiratory symptoms; nearly 60% frequency of comorbidities (diabetes, hypertension, and ischemic heart disease); and low rate of successful revascularization compared with that of usual thromboembolic limb ischemia.

## INTRODUCTION

Coronavirus Disease 2019 (COVID-19), causing an acute respiratory syndrome due to infection with coronavirus 2 (SARS-CoV-2), has affected Pakistan, as it has many other countries in the world; nationwide spread of COVID-19 sparked soon after the first reported case in February, 2020, and the disease has infected over half a million people and caused over 10,500 deaths in Pakistan.

COVID-19 infection has been reported to predispose to arterial and venous thrombosis leading to deleterious events. ^1^ Venous thrombosis is being increasingly reported, but data on arterial thrombosis, which may cause serious clinical challenges to vascular specialists, are sparse. SARS-CoV-2 virus enters human cells by latching its spikes onto the angiotensin converting enzyme 2 receptors on the surface of cells in the respiratory tract, blood vessels, and other sites. Vascular thrombosis may be mediated partially by coagulopathy, as demonstrated by elevations in D-dimer and fibrinogen levels.^2^ Acute thromboembolic phenomena seem to be especially frequent in COVID-19 patients, which has led to becoming a mainstay in management of COVID inpatients.^3^

We report here a series of 44 COVID-19 patients who presented with clinical, radiological, and laboratory evidence of arterial thrombosis causing limb-threatening ischemia.

### Patients and Methods

This was a nested case series using data from the prospectively maintained database of the vascular department of the University Hospital at Islamabad, Pakistan. This unit receives referrals from neighboring cities in the north of Pakistan. Forty-four patients suffering from mild to severe COVID-19 infection and presenting with critical limb ischemia were diagnosed and managed between March 2020 and December 2020. Seven patients, who presented with bluish discoloration of the hands or feet but with preserved peripheral pulses are not included in this series. Demographics of the patients are given in Table 1. Most patients (72.7%). were males, and 40.9% had no underlying co-morbidities; others had combinations of diabetes, hypertension, ischemic heart disease, and atrial fibrillation, and 29.6% were on an antiplatelet drug prior to their diagnosis of COVID.

**Table 1.**
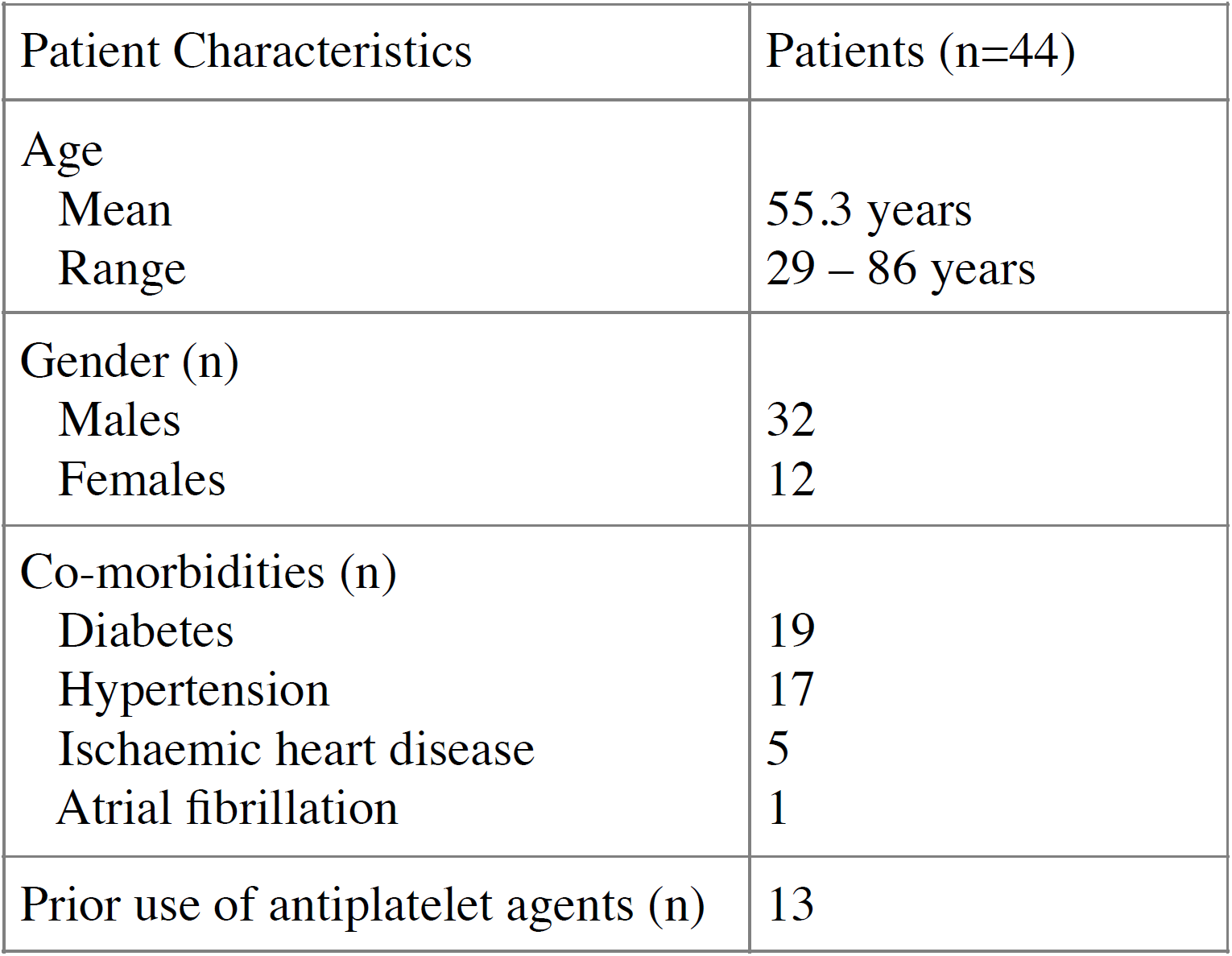

The diagnosis of COVID-19 was based on positive real-time reverse transcription–polymerase chain reaction (RT-PCR), serology (anti-SARS COV-2 antibodies), and high-resolution chest CT. Limb vasculature was evaluated through clinical assessment, CT angiography, and duplex scans. Management of the patients included anticoagulation with low molecular weight heparin (enoxaparin) followed by rivaroxaban, revascularization by thrombo-embolectomy or bypass grafting, and amputation. Based on the mode of presentation, the patients were divided into three groups: 1) already admitted to hospital, 2) presenting to accident and emergency department, and 3) presenting to vascular clinics.

## RESULTS

### Patients already admitted to designated COVID-19 area

Eleven patients (25%), who were already admitted in the designated COVID-19 area of the hospital for moderate to severe COVID-19 infection, presented with critical limb ischemia, and the vascular team was consulted **(Table 2)**. Eight of the 11 (73%) were on critical support with severe COVID-19 pneumonia and its complications. The other 3, with moderate respiratory syndrome, were managed in the ward. The 11 patients developed ischemic symptoms while they were receiving heparin as part of their treatment for COVID; they all had high D-dimer values (mean 13972), and 2 had deep venous thrombosis also. Nine of the 11 (82%) patients had preexisting comorbidities.

**Table 2.** Inpatients

|  |  |
| --- | --- |
| Patients admitted with COVID-19 | 11 |
| Comorbidities | 9/11 |
| Diabetes | 7 |
| Hypertension | 5 |
| Ischaemic heart disease | 3 |
| Atrial fibrillation | 1 |
| Prior use of antiplatelet agents | 6 |
| D-dimer (ng/ml) |  |
| Mean | 13972 |
| Range | 2400 - 20000 |
| Severity of COVID |  |
| Severe | 8 |
| Moderate | 3 |
| Limb involved |  |
| Lower limb | 9 |
| Unilateral | 8 |
| Bilateral | 1 |
| Upper limb | 2 |
| Proximal location of thrombosis |  |
| Aortoiliac | 2 |
| Femoropopliteal | 6 |
| Tibial and pedal | 1 |
| Brachial (with mesenteric) | 2 |
| Outcome |  |
| Death | 4 |
| Major amputation | 0 |
| Minor amputation | 4 |
| Limb salvage | 3 |

The limb ischemic events occurred between the 3^rd^ and 8^th^ days of onset of COVID symptoms in 9 patients and on the 20^th^ to 26^th^ day of onset of symptomatic infection in 2. Nine (82%) patients had lower-limb ischemia; 2 had ischemic upper limbs along with mesenteric ischemia.

Only 2 of the 8 critically sick patients were suitable for surgical revascularization. One had fresh thrombus at the aortic bifurcation extending bilaterally into the common iliac and right popliteal arteries; revascularization with limb salvage was accomplished with bilateral femoral embolectomy. The second patient had distal embolism from fresh thrombus in the iliac artery, which was removed; however, distal occlusion resulted in loss of toes.

Two patients had acute-on-chronic femoropopliteal occlusion diagnosed on CT angiogram. They were too ill for surgical revascularization, so they were managed with anticoagulation and required forefoot/toes amputation.

Two patients with ischemic leg had occlusion in the femoral and popliteal segments; they were too ill for any intervention and died. The patients with upper limb and mesenteric ischemia had extensive gangrene of the intestine seen on CT scan; they also died.

Three patients had moderate COVID infection, which was managed in the ward with high-flow oxygen, steroids, and heparin; one had unilateral femoropopliteal occlusion revascularized with femoral embolectomy; two had popliteal thrombosis with distal embolization into digital vessels; both were managed with anticoagulants and amputation of demarcated toes.

### Patients presenting to the emergency department

Seventeen patients presented with an ischemic limb through the emergency department (Table 3). Ten of the patients were asymptomatic/mild for COVID but with positive PCR. Seven had moderate symptoms; 3 were treated in hospital, and 4 were managed by the home outreach team, with regular monitoring, oxygen supplementation, and supportive care. One patient, who was asymptomatic at presentation, worsened post operatively and required intensive care with high-flow oxygen and continuous positive airway pressure. Five of the 17 patients (29%) had preexisting comorbidities and were getting single-agent antiplatelet (aspirin or clopidogrel) therapy. None of the 17 patients was receiving anticoagulants prior to reporting to the emergency department.

**Table 3.** Patients presenting through the emergency department

|  |  |
| --- | --- |
| **Total patients** | **17** |
| Comorbidities | 5/17 |
| Diabetes | 2 |
| Hypertension | 4 |
| Ischaemic heart disease | 1 |
| Prior use of antiplatelet agents | 5 |
| D-dimer (ng/ml) – 5 patients | 1620 - 18200 |
| Severity of COVID before limb ischaemia |  |
| Asymptomatic/mild | 10 |
| Moderate | 7 |
| Severe | 0 |
| Limb involved |  |
| Lower limb | 10 |
| Upper limb | 7 |
| Proximal location of thrombosis |  |
| Aortoiliac | 2 |
| Femoropopliteal | 5 |
| Subclavian | 5 |
| Brachial | 5 |
| Outcome |  |
| Death | 0 |
| Major amputation | 4 |
| Minor amputation | 0 |
| Limb salvage | 13 |

Ten (59%) patients who presented to the emergency room had upper-limb critical ischemia, and 7 (41%) had lower-limb critical ischemia. The peripheral ischemic event in these patients occurred between 3 days and 9 weeks (mean 15.6 days) after onset of COVID symptoms or history of exposure with positive PCR.

Of the 10 patients who were asymptomatic or had mild symptoms, 8 presented early, within 10 hours (mean 7.2 hours) of onset of symptoms. These patients had successful surgical embolectomy (6 brachial and 2 femoral), followed by prolonged anticoagulation. Two with subclavian-axillary thrombosis had motor and sensory impairments on arrival. The limbs were saved through revascularization; however, one patient developed wrist drop. In 6 patients, the acute thrombus was in the subclavian or brachial artery, with distal emboli, and 2 patients had iliofemoral thrombosis; 1 of these patients required a redo procedure within 24 hours.

Two patients presented to the emergency department later than 24 hours after onset of ischemic symptoms. One presented at 30 hours with thrombus in the distal subclavian and proximal axillary artery causing severe hand ischemia, which was successfully revascularized with brachial embolectomy and anticoagulation with no residual deficit. The other patient, who presented at 72 hours after onset of ischemic symptoms, had popliteal artery thrombosis with advanced ischemia that required a below-knee amputation.

The 7 patients who were moderately symptomatic for COVID-19 respiratory infection presented with ischemic limb after complete recovery from the COVID symptoms. Only one of these presented early--within 4 hours of onset of symptoms; he had successful brachial embolectomy. Three patients presented between 3 and 5 days after recovery from COVID symptoms with established signs of ischemia and early demarcation requiring major amputation (2 below-knee and 1 mid-forearm amputation). Three other COVID-symptomatic patients presented within 2-4 days of onset of ischemic symptoms (1 upper limb and 2 lower limb) but had no motor or sensory loss; CT angiography in them revealed occlusion in the subclavian artery and femoropopliteal segment, respectively; they recovered well with anticoagulation, except for mild claudication at 3 months follow-up (CT scan image). (CT images)

### Patients presenting to the outpatient department

Sixteen patients presented to vascular clinics with symptoms of ischemic limb (Table 4). Of these, 12 (75%) had comorbidities; only 5 of the 16 were receiving long-term antiplatelet drugs. Eleven patients had histories of mild COVID infection with no prior hospital contact. Five patients had moderate to severe COVID symptoms and were hospitalized for 1-3 weeks. All 16 patients developed limb ischemia after discharge from the hospital. The diagnosis of COVID was made by a positive PCR result in 13 patients, while 2 had coronavirus antibodies, and 1 had typical COVID-related changes on chest CT. D-dimer result was available for 1 patient only.

**Table 4.** Patients presenting through the outpatient department

|  |  |
| --- | --- |
| Total patients | 16 |
| Comorbidities | 12/16 |
| Diabetes | 10 |
| Hypertension | 6 |
| Ischaemic heart disease | 2 |
| Prior use of antiplatelet agents | 5 |
| D-dimer (ng/ml) – 1 patient | 15600 ra |
| Severity of COVID before limb ischaemia |  |
| Asymptomatic / Mild | 11 |
| Moderate | 5 |
| Severe | 0 |
| Limb involved |  |
| Lower limb | 13 |
| Upper limb | 2 |
| Both upper and lower limb | 1 |
| Proximal location of thrombosis |  |
| Aortoiliac | 2 |
| Femoropopliteal | 10 |
| Tibial | 2 |
| Subclavian | 0 |
| Brachial | 2 |
| Outcome |  |
| Death | 0 |
| Major Amputation | 6 |
| Minor Amputation | 2 |
| Limb Salvage | 10 |
| Anticoagulation only | 7 |
| Femoropopliteal bypass | 2 |
| Popliteal thromboembolectomy | 1 |

Thirteen of the 16 patients had lower limb ischemia, 2 had upper limb ischemia, and 1 had symptoms in both the upper and lower limbs. The average time for developing ischemic symptoms after onset of COVID-19 was 12.8 (5-30) days.

Of the five patients with moderate to severe COVID infection, 3 presented with irreversible ischemia and required major amputations. The other 2 had viable limbs with ischemic toes; they had revascularization with femoropopliteal bypass requiring only minor amputations.

Of the 11 patients with mild COVID symptoms, 3 presented with irreversible ischemic changes and had major amputations. Six of them, who had mild to moderate ischemic symptoms with viable limb (4 lower limbs and 2 upper limbs), were managed with anticoagulation; they have had reasonable symptomatic relief and are under surveillance. Two patients presented with pain at rest and blue toe syndrome. In 1, CT angiography revealed superficial femoral-popliteal thrombus with free-floating edges; the patient had popliteal thrombo-embolectomy with complete recovery. The other patient had proximal aortic thrombus with mild symptoms in the hand and gangrenous little toe, which was managed with anticoagulation and toe amputation.

The flow charts 1 and 2 give the outcomes as per patient presentation and severity of COVID-19 disease.

## DISCUSSION

This series of 44 COVID-19 patients highlighted the oft underappreciated seriousness and complexity of presentation, diagnosis, and management of arterial thrombosis causing limb-threatening ischemia in this disease. Whereas the association of venous thrombosis with COVID-19 has been reported repeatedly, the occurrence of acute limb ischemia in patients with COVID-19 is a newly recognized condition.

Several important clinical features of acute limb ischemia associated with COVID-19 were brought to light in this series: It is a vascular emergency and if not managed in a timely manner can result in limb loss and even death; 10 major amputations and 4 deaths occurred in this small cohort. The initial expression of ischemia may be subtle and ignored by patients and physicians, and the severity of respiratory infection and other symptoms may be inconsistent with the severity and level of vascular involvement; of 17 patients who presented through the emergency room with limb-threatening ischemia, 10 were asymptomatic for respiratory and general symptoms, and 7 were only moderately sick. Comorbidities were common among patients admitted to hospital as well as those seen in the emergency department or in clinics (26/44; 59%); the commonest were diabetes mellitus, hypertension, and ischemic heart disease. Limb ischemia was more common in males (72%), and persons as young as 29 years of age were affected. Anticoagulation did not always prevent the thromboembolic events, as all admitted patients were receiving low molecular weight heparin when they developed peripheral arterial thrombosis.

The rate of successful revascularization was deplorably low in COVID-19-related ischemia compared to that of routine patients of similar category, and distal limb (fore foot and distal phalanx) was affected despite revascularization attempts. This difficulty could be partially related to failure of distal revascularization and recurrent thrombo-embolic phenomena. Revascularization occurred in a few patients who were treated with anticoagulation therapy (continuous heparin infusion) only; this result may have been due to binding of the pathogens to the heparin rather than to the endothelial cells of the organism by inhibiting cell penetration.^21^ In our experience, with adequate knowledge, clinical suspicion of limb ischemia, attentive monitoring, and timely therapeutic measures for thrombotic complication of COVID-19 may end with fortunate outcome. Indeed, limbs were salvaged in 26/44 patients (59%), although the salvage rate among inpatients (3/11; 27%) was much lower than among patients seen in the emergency department (3/17; 76%) or in clinics (10/16; 62%).

The incidence, etiological factors, pathophysiology, and phenotype of acute limb ischemia in COVID-19 are still under investigation. SARS-CoV-2, like other members of the coronavirus family, causes respiratory infection, but in some patients, the infection triggers inappropriate immune responses, resulting in release of cytokines and chemokines leading to multi-organ and system impairment.^22^ Vascular damage is mediated by endothelial injury, coagulopathy, and hypercoagulability leading to thromboembolic endothelial cell injury; hyperimmune reaction, and platelet aggregations are blamed as initiating factors for thrombosis leading to ischemia.^23^ A common etiology of acute limb ischemia other than preexisting peripheral arterial disease is embolization.^16^ However, in patients with COVID-19, due to the nature of native thrombi in the vessels,^17,18^ it is thought that the virus induces a hypercoagulable state. Though myocardial injury and arrythmias caused by COVID-19 infection may lead to thromboembolic phenomenon, direct vascular endothelial injury or endothelial inflammation due to endothelial-cell infection has also been proposed.^6–8^

The common pattern of coagulopathy in COVID-19 infection is raised D-dimer and fibrinogen levels. Fibrin stimulates profibrotic growth factors, which in turn damage the vascular endothelium, the integrity of which is considered a barrier to clot formation and propagation.^9^ Complement activation also causes endothelial cell injury, leading to cell death and exposure of the thrombogenic basement membrane; more than 70% of patients who died of COVID-19 reportedly had DIC10.

Thromboembolism is also mediated by cytokines and chemokines, thus intensifying the vicious cycle of thrombosis leading to acute vascular insufficiency.^11^ COVID patients in critical care have been reported to have imbalance of hemostasis, with decreased platelet count, low fibrinogen, and increased D-dimer, consistent with a hypercoagulable state. Factors that contribute to coagulation, such as Factor VIII, vWF, vWF antigen, have been found increased in many patients. Antiphospholipid antibodies also have been found, although their link with thromboembolic events remains unclear. ^12^

Monitoring D-dimer and fibrin degradation products values may be helpful for the early identification of severe COVID-associated thromboembolism. ^13, 14^ In our series, data on D-dimer values were incomplete; the patients who presented in clinics late with limb ischemia had no recorded D-dimer values, whereas admitted patients in acute phase of disease had D-dimers measured, which were found elevated.

The incidence of thromboembolic events in COVID patients, causing life threatening conditions, might be higher than reported.^19^ A study conducted in Italy revealed a direct increase in incidence of acute limb ischemia related to COVID, deduced from a significant increase in numbers during the pandemic months compared with the same time in the previous years.^20^

This study has limitations. It is a single-site study of modest-size population; studies of larger populations in multiple sites are needed to assess its general applicability. It is a retrospective study, albeit of prospective patient accrual, so it has inherent potential for biases.

## CONCLUSION

The occurrence of acute limb ischemia in patients with COVID-19 is a newly appreciated condition. The infection leads to inflammation and a prothrombotic condition resulting in thrombosis of the vessels and ischemia of various degree depending upon the level and nature of the vessel involved. It is a vascular emergency that if not managed in a timely manner can result in limb loss and even death. Clinical awareness of the subtle expression of acute limb ischemia in COVID-19 disease and prompt therapeutic intervention are needed. The pathophysiology of acute limb ischemia in COVID-19 is poorly understood and requires thorough study.

**Flow Chart 1:**
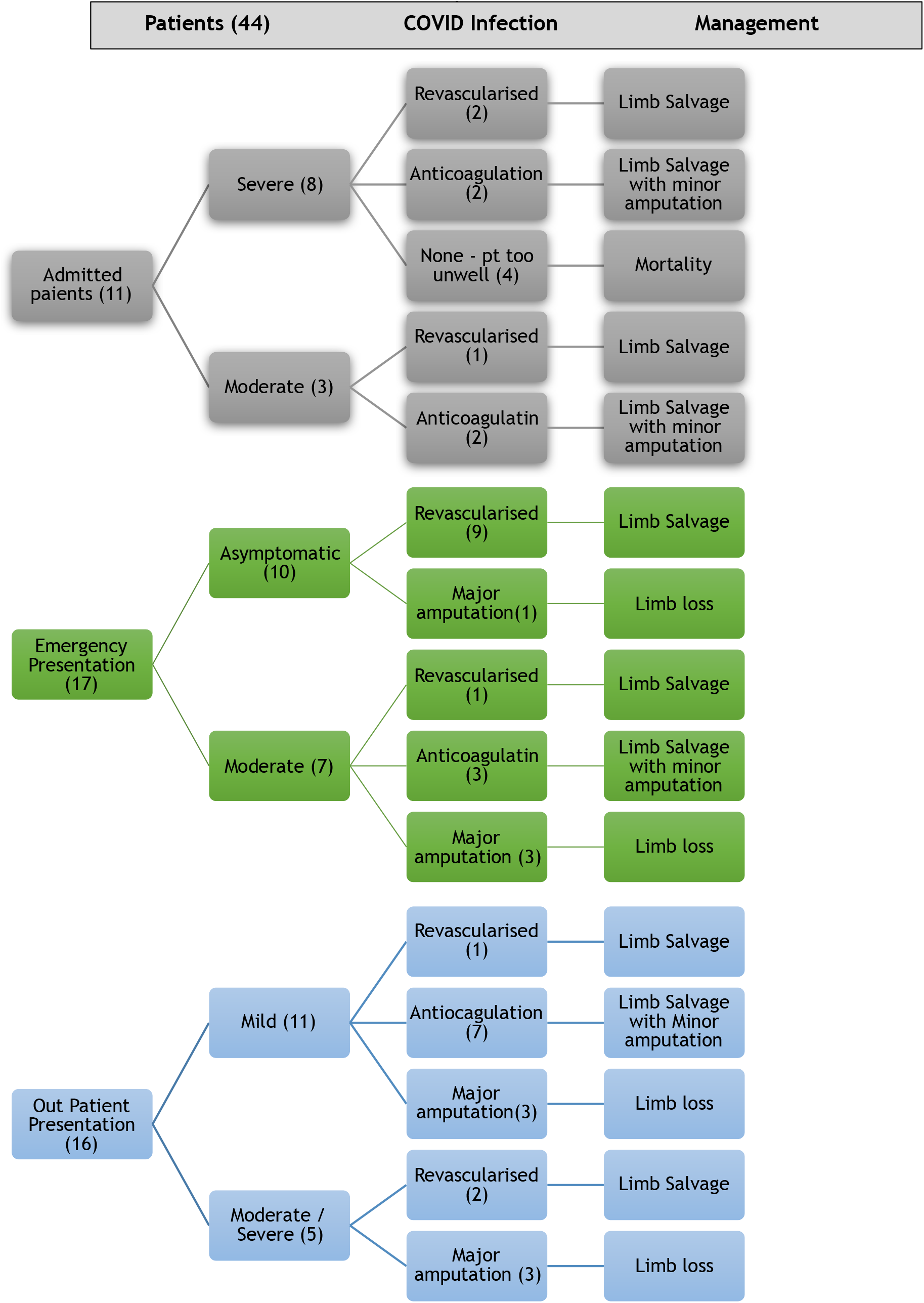
Patients presentation and their outcomes.

**Flow Chart 2:**
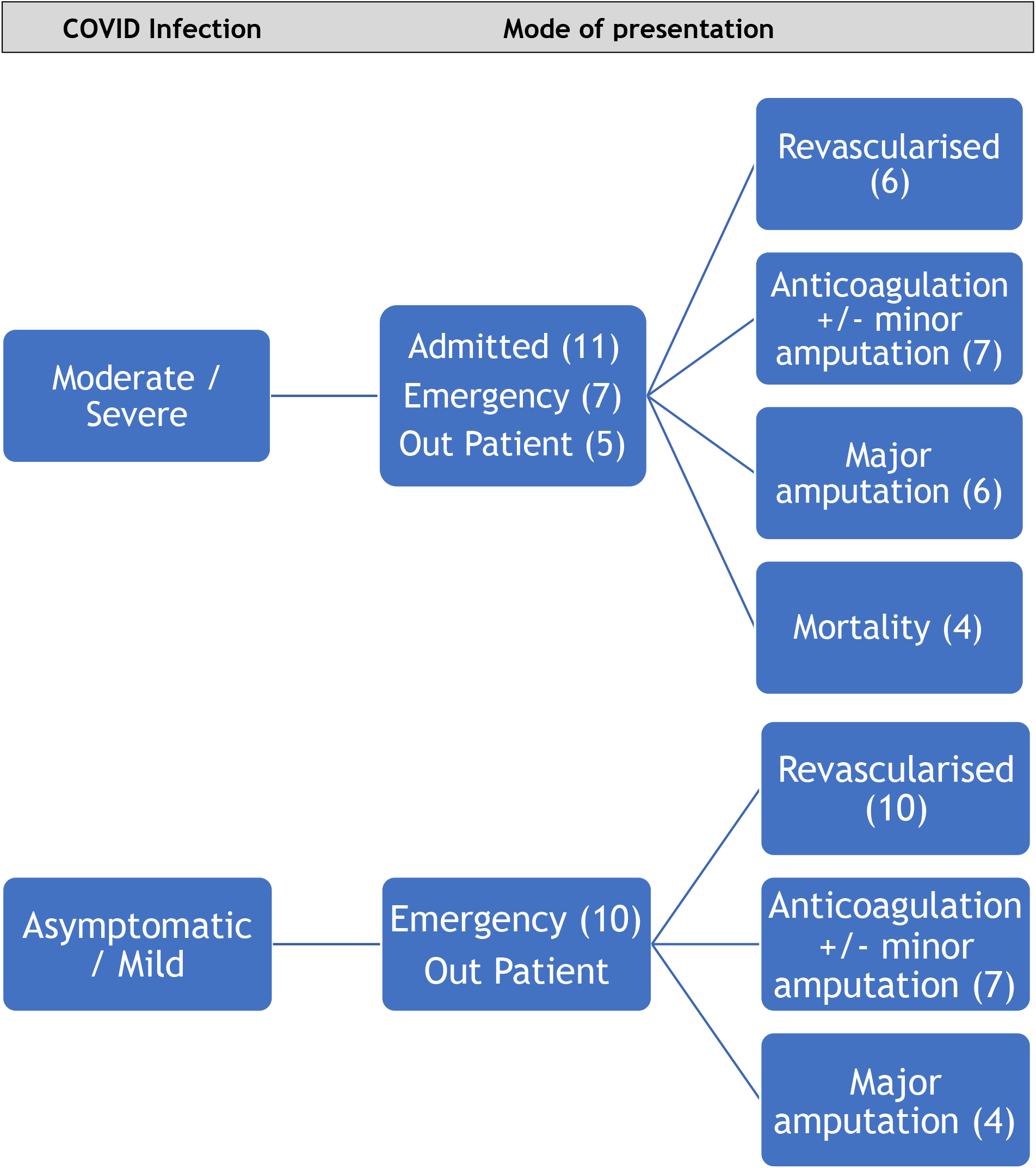
Severity of COVID-19 infection and outcomes

## Supporting information

IRB & Ethics committee approval

## Data Availability

data will be avialiable on request

---

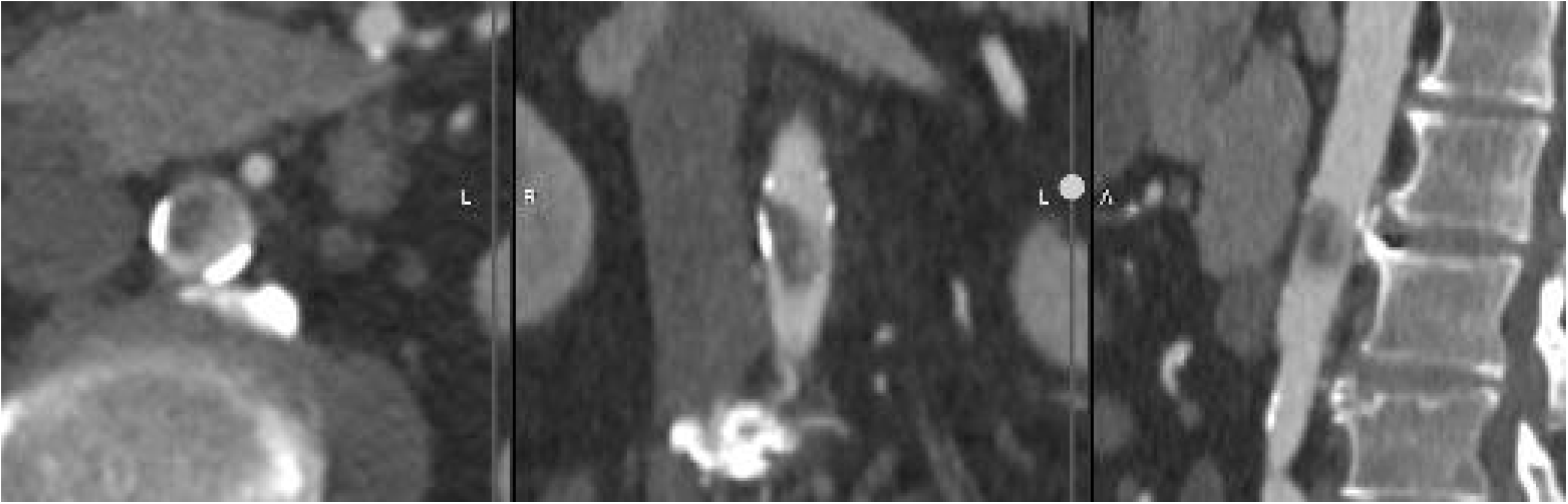

