## Supplementary material for "Management of COVID-19-related Arterial Thrombosis Leading to Acute Limb-threatening Ischemia": IRB & Ethics committee approval

### Institutional review board and Ethics committee-IRB/EC

Rawalpindi Medical University
District Headquarter Hospital Rawalpindi

July 02, 2020

Dr Mohammad Waqas Raza
DHQ hospital Rawalpindi

Ref: IRB02-8765

Dear doctor

Please note that with reference to your study "Management of COVID-19-related Arterial Thrombosis Leading to Acute Limb-threatening Ischemia" has been reviewed by the IRB/EC. The IRB & Ethic committee is pleased to approve the study.

The IRB/EC is in accordance with the ICH and GCP guidelines. Any changes in the protocol should be notified to the committee for prior approval. All the informed consents should be retained for future reference. An updated report should be submitted to the IRB & Ethics Committee.

Sincerely,

**Prof Iqbal Khan**
Chairman, IRB/EC
